# Liquid Biopsy for Detection of Pancreaticobiliary Cancers in Suspected Patients by Functional Enrichment and Immunofluorescent Profiling of Circulating Tumor Cells and their Clusters

**DOI:** 10.1101/2024.02.13.24302805

**Authors:** Andrew Gaya, Nitesh Rohatgi, Sewanti Limaye, Aditya Shreenivas, Ramin Ajami, Dadasaheb Akolkar, Vineet Datta, Ajay Srinivasan, Darshana Patil

## Abstract

**Background:** Circulating tumor cells (CTCs) have historically been used for prognostication in oncology. We evaluate performance of liquid biopsy CTC assay as a diagnostic tool in suspected pancreaticobiliary cancers. The assay utilises functional enrichment of CTCs followed by immunofluorescent profiling of organ specific markers.

**Methods:** Multicentric case control study was followed by a prospective observational study. Adult patients undergoing tissue sampling for suspected pancreaticobiliary cancer were included in the studies. Blood samples for TruBlood® CTC assay were drawn before tissue sampling. Patients with a prior cancer treatment or cancer history were excluded. CTCs and their clusters were harvested by a unique functional enrichment method which is label-free, size independent and non-mechanical. The CTCs then underwent immunofluorescent profiling for CA19.9, Maspin, EpCAM, CK and CD45, blinded to the tissue histopathological diagnosis. TruBlood® malignant or non-malignant predictions were compared with tissue diagnoses to establish the sensitivity and specificity.

**Results:** The case-control study evaluated 360 participants including 188 cases diagnosed with pancreaticobiliary cancer and 172 healthy individuals. A subsequent prospective observational study included 88 individuals with suspicion of pancreaticobiliary malignancy. The test had 95.9% overall sensitivity (95% CI: 86.0% - 99.5%) and 92.3% specificity (95% CI: 79.13% to 98.38%) to differentiate pancreaticobiliary cancers (PBC) (n = 49) from benign PB conditions (n = 39).

**Conclusions:** The high accuracy of the CTC based TruBlood test demonstrates its potential clinical application as a diagnostic tool to assist effective detection of pancreaticobiliary cancers when tissue sampling is unviable or inconclusive. A confirmational prospective interventional studiy in patients with suspicion of pancreaticobiliary malignancy including those with unavailability of tissue diagnosis is warranted.

## BACKGROUND

Early-stage cancers have better survival outcomes than late-stage disease [1]. This gap is exaggerated in aggressive poor prognosis malignancies like pancreaticobiliary cancers (PBC) [2–4]. The early stages of PBC are generally asymptomatic, whilst advanced cases tend to have non-specific symptoms such as abdominal pain, anorexia, and weight loss. Most PBC are detected at advanced stages (Stage IV) where 5-year survival rates are <5% [2–4]. Globally, there are no recommendations for PBC screening in non high-risk individuals. Individuals at higher risk for pancreatic cancers may be advised endoscopic ultrasound (EUS) based or computed tomography (CT) surveillance [5]. The relatively low incidence of PBC may imply a lower benefit ratio from screening large populations for PBC. The United States Preventive Services Task Force (USPSTF) guideline cites the unavailability of accurate tests for detection of PBC [6]. There is thus a large unmet clinical need. This clinical challenge is amplified by diagnostic challenges in a subset of patients. The diagnostic work-up for suspected cases of PBC includes evaluation of serum cancer antigen 19-9 (CA19-9) and carcinoembryonic antigen (CEA), followed by medical imaging such as EUS, CT, endoscopic retrograde cholangiopancreatography (ERCP) or magnetic resonance cholangiopancreatography (MRCP), during which tissue sampling is also performed for histopathological examination (HPE) to establish a conclusive diagnosis [7,8]. Diagnostic challenges include the inability to obtain (sufficient) tissue samples due to inaccessible location of the tumor, as well as incomplete or inconclusive HPE findings due to non-diagnostic tissue which may be seen in up to 11% of suspected cases [9–13]. Further, about 31% of cases with negative HPE findings have been reported to be false negatives [9–12]. EUS guided biopsy rather than fine needle aspiration cytology (FNAC) has improved the diagnostic yield, but is invasive and not without significant risks. These challenges delay diagnosis and time to treatment due to resource constraints. The standalone evaluation of serum antigens has low sensitivity as well as specificity in the symptomatic population and thus is not relied upon as a diagnostic aid.

A non-invasive test with high sensitivity and specificity for detection of PBC is an unmet clinical need which could also have potential for PBC screening. Evaluation of circulating tumor cells (CTC) [14] has the potential to facilitate more effective detection of PBC. CTCs are ubiquitously detected in solid tumors and are highly specific for malignancy. They are undetectable in individuals with non-malignant conditions such as benign or inflammatory conditions (pancreatitis or IgG4 disease) which may represent a diagnostic dilemma on imaging or serum tumor marker profiling. In prior studies, CTCs have been observed to retain the molecular characteristic features of the primary malignancy, and it is proposed that their evaluation can provide non-invasively diagnostically relevant information on the underlying malignancy [15].

We describe a non-invasive liquid biopsy test (TruBlood®) which enriches and detects PBC associated circulating tumor cells (CTCs) and their clusters from peripheral blood samples. This test is based on our previously described functional enrichment process to isolate CTCs as single cells as well as in clusters [16], which can be profiled by immunocytochemistry (ICC) to obtain diagnostically relevant information on underlying malignancy [17]. In suspected PBC cases, CTCs enriched from blood samples are evaluated by ICC to determine the expression of cancer antigen 19.9 (CA19.9), mammary serine proteinase inhibitor (Maspin) along with epithelial cell adhesion molecule (EpCAM), cytokeratins (CK), and the common leucocyte antigen (CD45). Positive Expression of CK and EpCAM and negative CD45 status confirm the epithelial origin of the CTC, while positivity for CA19.9 and / or Maspin suggest pancreaticobiliary tract origin. In the present manuscript, we present the clinical performance characteristics of the TruBlood® test.

## METHODS

### Study Participants and Samples

Biological samples mentioned in this manuscript were obtained from participants in three prospective observational studies, all of which are registered at Clinical Trials Registry - India (https://ctri.nic.in/Clinicaltrials/). The TRUEBLOOD study (Trial ID CTRI/2019/03/017918, ongoing) enrols patients diagnosed with or suspected of cancers and patients with benign (non-malignant) conditions. The CTC-based Cancer Detection and Diagnosis Study: (Trial ID CTRI/2022/02/040373, Feb 2022 - ongoing) enrols suspected cancer cases. The RESOLUTE study (Trial ID CTRI/2019/01/017219, Jan 2019 – ongoing) enrols healthy asymptomatic adults with no prior diagnosis of cancer. All studies were previously approved by the Ethics Committees (EC) of the sponsor (Datar Cancer Genetics, DCG) as well as of the participating institute(s). All studies are conducted in accordance with the Declaration of Helsinki.

Written informed consent was obtained from all study participants prior to collection of peripheral blood sample by venous draw into EDTA vacutainers. Leftover blood samples from patients who had availed of the sponsor’s services were also used (after obtaining EC approval and informed consent, in each case). Blood samples from suspected cancer cases were collected prior to the tissue sampling. All sample identities were masked with unique 10-digit alphanumeric codes, stored at 2°C −8°C and processed at the CAP and CLIA accredited facilities of the study sponsor, which also adhere to quality standards ISO 9001:2015, ISO 27001:2013 and ISO 15189:2012. The reporting of observational studies in this manuscript is compliant with STROBE guidelines [18].

### Markers, Antisera and Cell Lines

Details of the markers employed by the test as well as the antisera and cell lines used for method development or as controls are provided in Supplementary Methods (SM)1 and SM2. The purity of all cell lines used in the study was confirmed by periodic Short Tandem Repeat (STR) profiling. All cell lines were also periodically tested and verified to be Mycoplasma negativity.

### Enrichment of Circulating Tumor Cells from Peripheral Blood

Blood samples were processed for the enrichment of CTCs from white blood cells (WBCs) as described previously [16]. Briefly, WBCs were isolated from blood samples following lysis of red blood cells (RBCs) and then treated with a proprietary CTC enrichment medium (CEM), which induces cell death in all non-malignant cells while apoptosis reluctant malignant cells (CTCs) survive. After CEM treatment, surviving cells are retrieved for further use. The process is also explained in SM3.

### Immunocytochemistry Profiling of CTCs

The process of ICC profiling of CTCs was as described previously [17] and is also provided in SM4. Briefly, the CTCs are seeded into wells of an imaging compatible multi-well plate, fixed and sequentially immunostained with cocktails of fluorophore-conjugated antibodies (Ab) against the markers. Marker positive cells are then visualized using an appropriate fluorescence imaging system. A schema showing the various steps of the process including CTC detection and ICC profiling is depicted in **Figure 1**. Test samples received a Positive classification based on detection of CK+, CA19.9+, CD45-cells and / or CK+, Maspin+, CD45-cells, with or without detection of CK+, EpCAM+, CD45-cells. Samples with any other findings received a Negative classification.

**Figure 1.**
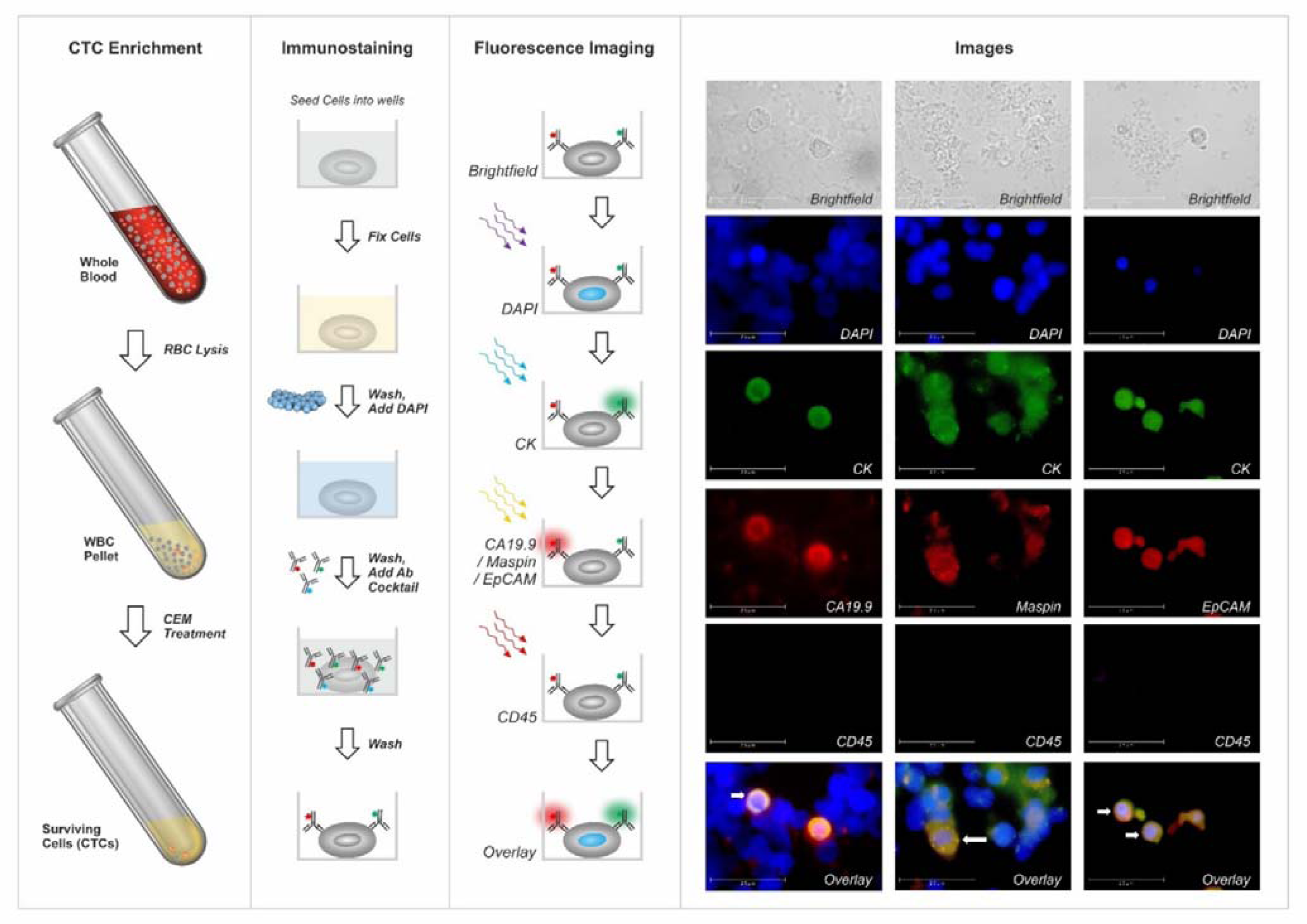
Schema of Test. White blood cells (WBCs) are isolated from blood samples following lysis of red blood cells (RBCs). Circulating tumor cells (CTCs) are then enrichched from WBCs using a CTC enrichment medium (CEM) that eliminates all non-malignant cells and permits tumor derived malignant cells to survive. The CTCs are seeded into imaging compatible multi-well plates and immunostained using antibody (Ab) cocktails to detect the status of CA19.9, Maspin, CK, EpCAM and CD45.

**Figure 2.**
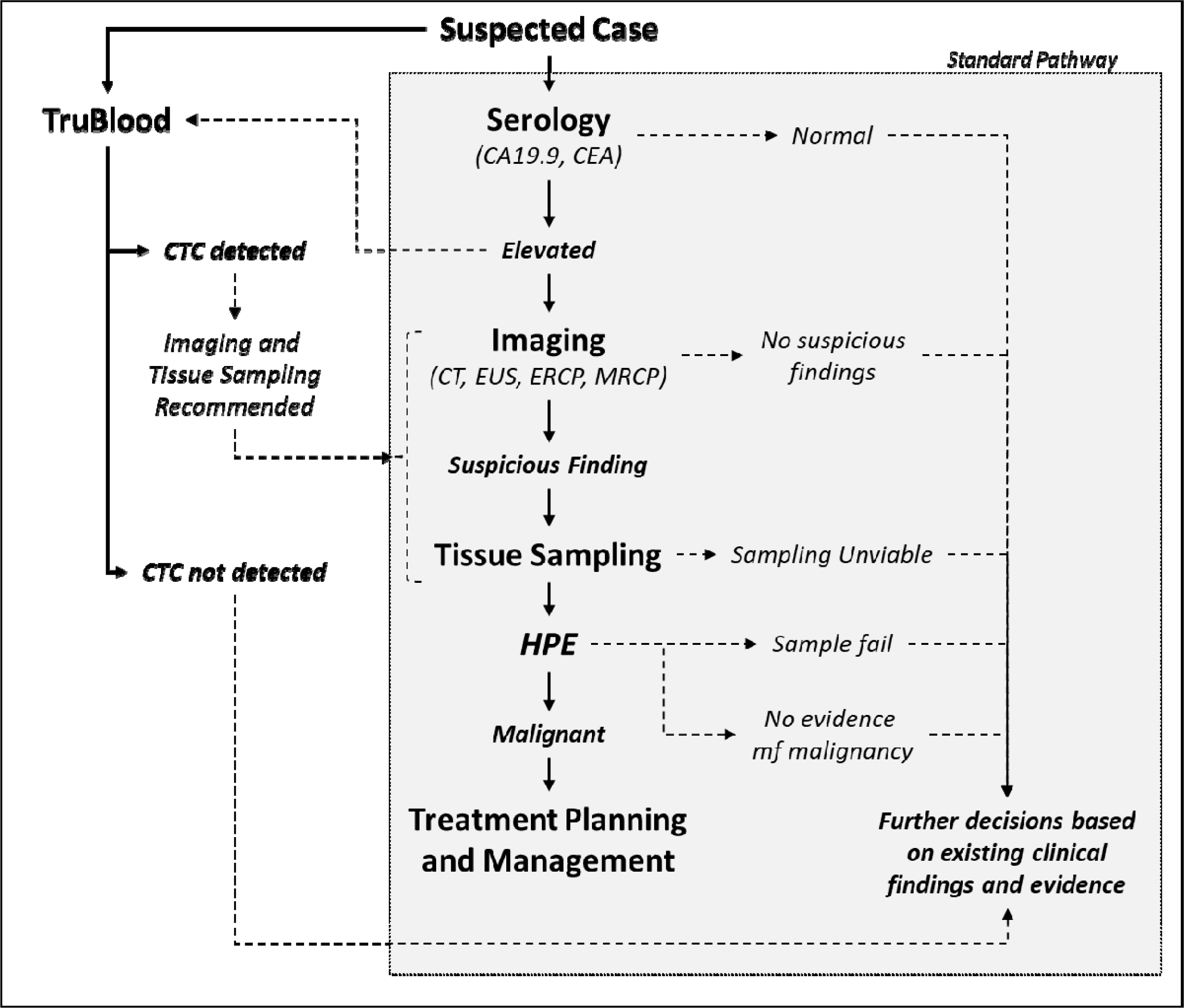
TruBlood Testing in the Standard Diagnostic Pathway. The TruBlood test described in the study is not intended to replace any of the tests in the standard diagnostic pathway, but to complement the same and provide additional evidence that may be interpreted in conjunction with clinical findings and evidence from the standard investigations. The potential advantage of the TruBlood test is to facilitate clinical decision making among those individuals where tissue based diagnosis is unavailable or unviable.

### Method Development and Validation

Comprehensive details of Method Development and Validation are provided in Supplementary Methods. These include determination of marker expression (marker specificity) in various cell types (SM5), impact of innate factors (age, gender, primary organ and stage) on marker expression (SM6), (non-)detectability of CTCs in non-malignant conditions (SM7), stability of CTCs in patient samples (SM8), sensitivity metrics of the test (SM9) including linearity and limit of detection, specificity metrics of the test including limit of blank (SM10) and interference (SM11) and inter-operator agreements (SM12).

### Case Control Clinical Study

The performance characteristics of the test to detect and differentiate PBC cases from asymptomatic individuals were ascertained and established in a case control study with samples from 360 participants, which included 188 recently diagnosed, therapy naïve cases of PBC and 172 asymptomatic adults, the latter having neither prior diagnosis of any cancer, no current suspicion of cancer, being generally asymptomatic, with normal findings on ultrasonography (USG) of the abdomen and normal serum CA19.9 levels. Samples of the asymptomatic cohort were randomized into Training and Test Sets in a 70%:30% ratio. The PBC cases were first segregated by Stage and then assigned to Training and Test Sets in the same ratio. All blood samples were processed for CTC enrichment and detection of marker (CK, EpCAM, CA19.9, Maspin and CD45) by operators unaware of the clinical status of the samples.

The marker expression status in the Training Set samples (n = 132 PBC, n = 120 asymptomatic) was correlated with their clinical status to ascertain marker positivity in cancer samples as well as absence of such marker positive cells in asymptomatic samples. Next, the marker expression status of the Test Set samples (n = 56 PBC, n = 52 asymptomatic) was used to predict the clinical status. The concordance of the prediction with the actual clinical status was used to determine the sensitivity (rate of true positives), specificity (rate of true negatives) and accuracy (combined rate of true positives and true negatives) of the case-control cohort test set.

Subsequently, all Training Set and Test Set samples (PBC and asymptomatic) were digitally pooled and random 30% samples from PBC (stage-wise) and asymptomatic patients were selected. The identities of these samples were re-masked and provided for the prediction of clinical status and the determination of sensitivity and specificity.

The above pooling-resampling-remasking-reanalysis was repeated successively to obtain 20 iterations of the Test Set including the original assignment. At each of these iterations, the sensitivity and specificity of the prediction were determined. The median sensitivity across the 20 iterations of the cross-validation process as well as the overall specificity and accuracy was reported along with the 95% confidence interval (CI) and the range.

### Prospective Clinical Study

The performance characteristics of the test to diagnose PBC and to differentiate it from benign PB conditions (PBB) were ascertained and established in a prospective clinical study with blood samples from 88 patients with no prior diagnosis of cancer and who presented with symptoms and radiological findings suspected of PBC. All patients underwent routine tumor tissue sampling for standard histopathological diagnosis. Pre-biopsy blood was collected from all patients and processed for CTC enrichment and detection of marker positive cells. Based on the marker expression status, the samples were classified as Positive or Negative. The clinical status (histopathological diagnosis: malignant v/s benign) of the samples was then unmasked to the sponsor to evaluate the concordance of the CTC-based prediction model with the clinical status (histopathological diagnosis) and determine the performance characteristics of the test.

## RESULTS

### Method Development and Validation

Summarized findings of the Method Development and Validation studies are provided in the respective Supplementary Methods sections along with indicated Supplementary Tables (ST)1-ST5 and Supplementary Figures (SF)1-SF4.

### Case Control Clinical Study

The performance characteristics of the test were evaluated in a case control study. The study inclusion criteria are provided in ST6, and the demographics of the study cohort are provided in ST7. The asymptomatic cohort was a South Asian population with <0.003% reported PBC incidence (in age > 40 years). Since the asymptomatic participants were also required to have normal CA19.9 and USG the risk of an underlying malignancy was further lowered. Samples in the Case Control Study were split into Training and Test set (70:30) and evaluated by a stringent, blinded, iterative manner which eliminated any risk of overfitting. The observations in the Training Set are provided in ST8.

In the absence of any positive findings among the asymptomatic samples (n = 52) in the Test Set, the specificity of the test (cancer v/s healthy) was 100% (95% CI: 93.15% - 100.00%). The median overall Sensitivity for detection of PBC (n = 56) was 96.4% (95% CI: 91.6% - 100%) and the overall Accuracy was 98.2% (95% CI: 93.47% to 99.77%). The number of samples (cancer-wise, stage-wise and overall), and median sensitivities (stage-wise and overall) (along with 95% CI and range across the 20 iterations) are provided in **Table 1**.

**Table 1.**
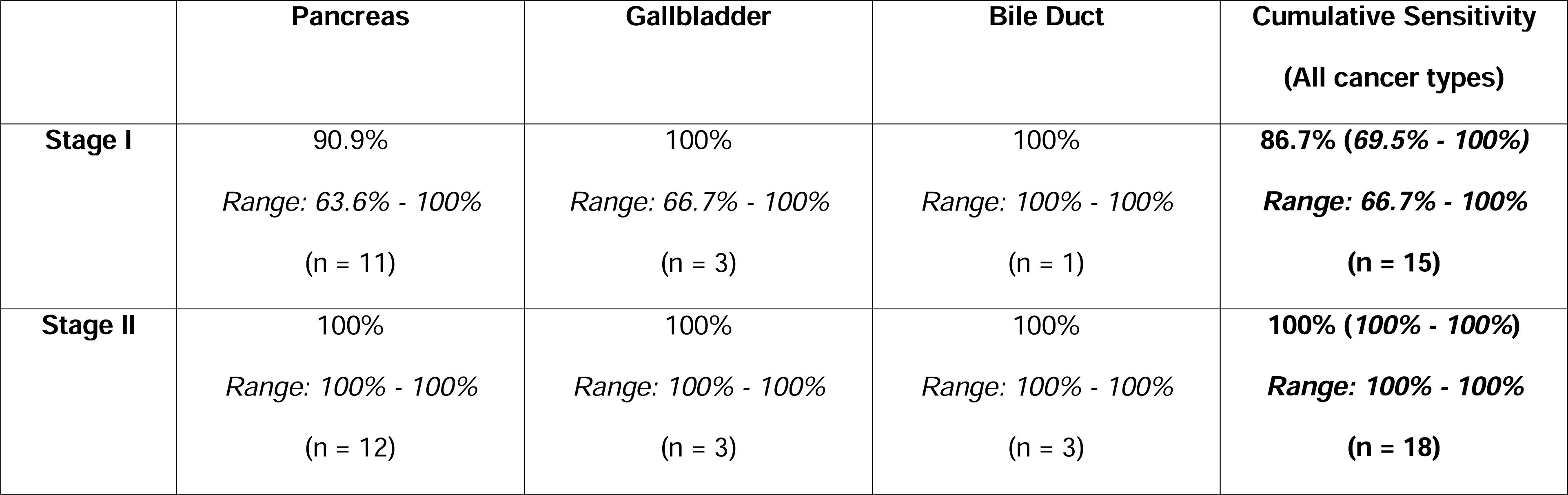
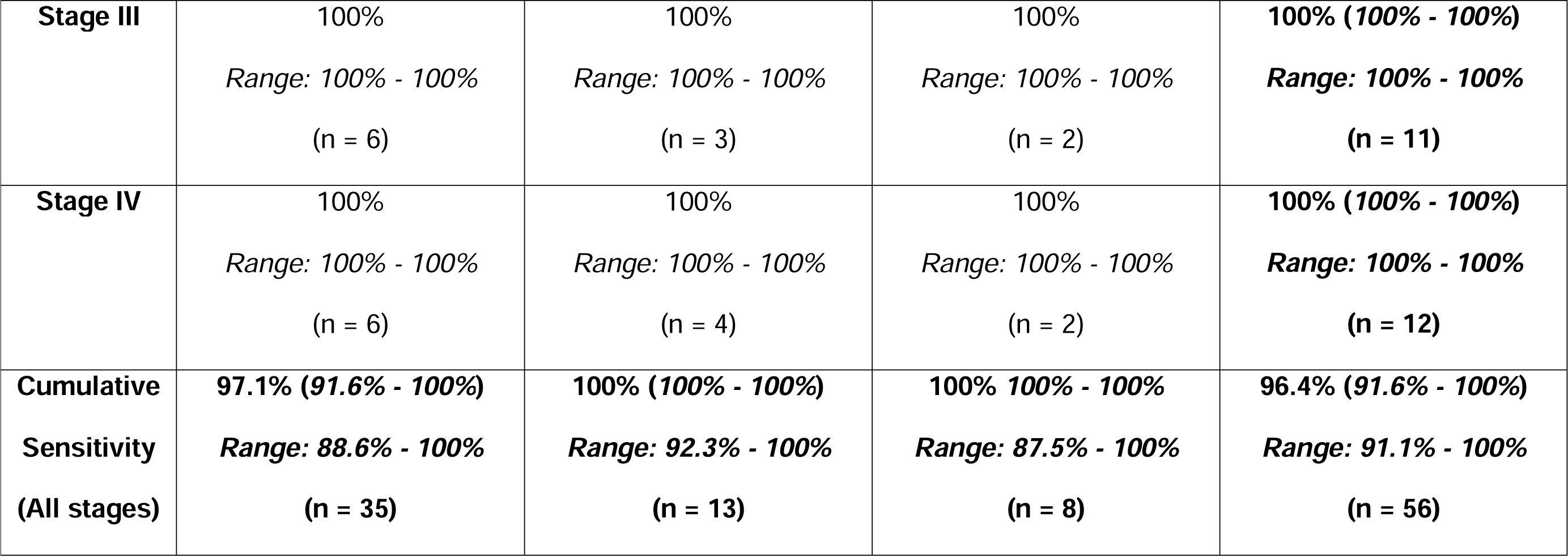
Sensitivity of the Test in the Case Control Study. Overall performance characteristics of the test as determined from 20 iterations of the Test Set. The table reports the median and range of stage-wise CTC detection rates, and the number of (n) pancreas, gallbladder and bile duct cancer samples, along with the cumulative (cancer-wise and stage-wise) Median Sensitivities. Values within parentheses adjacent to cumulative median sensitivities are the respective 95% Confidence Interval (95%CI) values.

### Prospective Clinical Study

The performance characteristics of the test to differentiate benign pancreaticobiliary conditions from malignant, were established in a second clinical study populated with 88 patients (45 females, 43 males, median age 51 years, age range: 21 - 81 years) presenting with clinical symptoms and radiological findings suspected of PBC. None of the patients had a prior diagnosis of cancer. Among the 88 patients, 39 were eventually diagnosed with PBB and 49 with PBC (AD). The demographic details of the study cohort are provided in ST9. The observations in the study samples are provided in ST10. The specificity of the test (cancer v/s benign) was 92.3% (95% CI: 79.13% - 98.38%). PB-CTCs were detected in 47 of 49 cancer samples yielding an overall clinical sensitivity of 95.9% (95% CI: 86.02% - 99.50%) and the overall accuracy was 94.32% (95%CI: 87.24% - 98.13%). The 2 false negative samples included a case of Stage 1 pancreatic cancer and a case of Stage 1 gallbladder cancer. The 3 false positive samples included a case of cholecystitis with cholelithiasis, a case of heterotopic pancreas and a case of cystic fibrosis of the pancreas. The cancer-wise and stage-wise and overall Sensitivities are provided in **Table 2**.

**Table 2.** Sensitivity of the Test in the Prospective Study. The table reports the CTC detection rates and stage-wise number of (n) pancreas, gallbladder and bile duct cancer samples, along with the cumulative (cancer-wise and stage-wise) sensitivities. Values within parentheses adjacent to the cumulative sensitivities are the respective 95% Confidence Interval (95%CI) values.

|  | **Pancreas** | **Gallbladder** | **Bile Duct** | **Cumulative Sensitivity**<br>**(All cancer types)** |
| --- | --- | --- | --- | --- |
| **Stage I** | 91.7%<br>(n = 12) | 88.9%<br>(n = 9) | - | **90.5% (69.6% – 98.8%)**<br>**(n = 21)** |
| **Stage II** | 100%<br>(n = 2) | 100%<br>(n = 2) | 100%<br>(n = 1) | **100% (47.8% - 100%)**<br>**(n = 5)** |
| **Stage III** | 100%<br>(n = 4) | 100%<br>(n = 2) | 100%<br>(n = 4) | **100% (69.2% - 100%)**<br>**(n = 10)** |
| **Stage IV** | 100%<br>(n = 5) | 100%<br>(n = 6) | 100%<br>(n = 2) | **100% (75.3% - 100%)**<br>**(n = 13)** |
| **Cumulative**<br>**Sensitivity** | **95.7% (78.1% - 99.9%)**<br>**(n = 23)** | **94.7% (73.9% - 99.9%)**<br>**(n = 19)** | **100.0% (59.0% - 100%)**<br>**(n = 7)** | **95.9% (86.0% - 99.5%)**<br>**(n = 49)** |
| (All stages) |  |  |  |  |

## DISCUSSION

We describe a liquid biopsy test for pancreaticobiliary cancer detection based on multiplexed fluorescence ICC profiling of CTCs functionally enriched from a blood sample. In two clinical studies, the test could detect adenocarcinomas (AD) of the pancreas, gallbladder and bile duct with high sensitivity (including for early stages) and differentiate PBC cases from healthy individuals, as well as individuals with benign conditions with high specificity, with a lower risk of false positives.

Our test is based on the detection of CTCs, which are ubiquitous in PBC and undetectable in individuals with benign conditions of the pancreas and biliary tract. Evaluation of CTCs can thus facilitate the detection of PBC with higher sensitivity as well as higher specificity due to relatively low risk of false positive findings. While prior studies have shown the presence of CTCs in pancreatic cancers [19–24], the technology platforms used in these studies are known to have lower specificity. These studies profile CTCs for disease prognostication and are limited in their ability to provide diagnostic profiling or screening.

The functional CTC enrichment and detection process of our test has distinct advantages over epitope capture which has lower sensitivity to capture and detect CTCs with low expression of target biomarkers (EpCAM and CK) as has been demonstrated in several prior studies [25–30]. This functional enrichment method can effectively detect CTCs as well as their clusters with no loss of sensitivity associated with age or gender of the patient or the primary or stage of disease. This advantage is reflected in prior clinical studies for this platform, where high CTC detection rates were consistently observed across all target cancer types and stages. The test also reported reliable performance in samples from asymptomatic individuals or those with benign conditions.

This CTC-based approach also has advantages over profiling of circulating tumor nucleic acids in blood sample, the latter having lower sensitivities, especially at the early stages, which hinder meaningful clinical applications [31].

The performance characteristics of the test has potential for dual utility of the technology platform, i.e., screening as well as diagnostic guidance / triaging. When used for screening, the test may be able to detect PBCs at earlier stages where the cancers may be more amenable for curative intent treatments. Detection of cancers at earlier stages combined with prompt treatment permits less aggressive treatment, leading to a better quality of life of the patient and is associated with significantly reduced mortality [32]. Early diagnosis can also significantly reduce the cost of treatment; treatment cost of early diagnosed patients was 25%-50% lower than for patients with advanced cancer [33]. Despite the lower incidence of PBC cancers, they are aggressive with poor outcomes, and have significant economic impact on the patients. Hence the asymptomatic / higher risk populations stand to benefit from screening and the associated potential for improved survival from early detection. The utility of this test to support diagnostic pathway is especially appreciable in suspected cases with comorbidities preventing tissue sampling, non-diagnostic tissue or inconclusive histopathological findings, as well as in patients with negative findings but clinical suspicion of malignancy. The test can minimize the dependence on tumor tissue sampling in suspected cases.

The high sensitivity and specificity of the test are its potential advantages. The test neither requires tumor tissue sample nor is it dependent on any foundational tumor tissue based test. Other compelling benefits of the test include its (a) safety, since the blood draw has low inherent procedural risks, (b) convenience, since the blood draw can be performed at any primary healthcare centre, (c) cost-effectiveness, since it requires no specialized facilities and can be integrated easily within existing clinical pathways, (d) low risk, since an inability of the test to perform as expected (i.e., test failure) does not deny the individual standard of care procedures or treatments, and, (e) versatility, being equally equipped to support detection of all PBC globally despite the relatively higher incidence of pancreatic cancers in the western hemisphere and relatively higher incidence of gallbladder cancers in the eastern hemisphere. The test is based on the detection of CTCs and localization to the pancreaticobiliary tract based on evidence of various cellular markers. There are currently no reports to suggest that the expression of these markers in cancers varies between various ethnicities. Thus race / ethnicity is not expected to be a confounding factor in the detection of PBCs. The test is currently not intended for detection of non(-adeno)-carcinoma subtypes such as neuroendocrine tumors which account for about 5% of PBC. However, future iterations of the test are envisaged to include additional markers for detection of these subtypes. Further, the test is currently not intended to distinguish between cancers of the pancreas, gallbladder and the bile duct. A limitation of our overall study is that it does not include a prospective study in the asymptomatic population to evaluate the clinical utility of the test for PBC screening. We propose such a study going forward. The strengths of our overall study include stringent analytical validation and clinical studies with (a) sample blinding to eliminate bias, (b) an iterative case-control design to eliminate risk of over-fitting and (c) prospective assessment of performance in suspected PBC cases.

## CONCLUSIONS

The study establishes the feasibility of liquid biopsy for detection of pancreaticobiliary cancers based on enrichment and characterization of circulating tumor cells (CTC). Further large(r) cohort studies in the respective intended use (IU) populations are planned to establish its clinical utility as a screening tool in high-risk individuals or for diagnostic guidance in suspected cases.

## ABBREVIATIONS

%: Percent
Ab: Antibodies
AD: Adenocarcinoma
CA19.9: Cancer Antigen 19.9
CAP: College of American Physicians
CD45: Common Leucocyte Antigen
CEA: Carcinoembryonic Antigen
CEM: CTC Enrichment Medium
CI: Confidence Interval
CK: Cytokeratin
CLIA: Clinical Laboratory Improvement Amendments
CT: Computed Tomography
CTC: Circulating Tumor Cell
ddPCR: Droplet Digtal Polymerase Chain Reaction
DCG: Datar Cancer Genetics
EC: Ethics Committee
EpCAM: Epithelial Cell Adhesion Molecule
ERCP: Endoscopic Retrograde Cholangiopancreatography
EUS: Endoscopic Ultrasound
FNAC: Fine Needle Aspiration Cytology
gDNA: Genomic DNA
HPE: Histopathological examination
ICC: Immunocytochemistry
IEC: Institutional Ethics Committee
IgG: Immunoglobulin G
LoB: Limit of Blank
LoD: Limit of Detection
MRCP: Magnetic Resonance Cholangiopancreatography
Maspin: Mammalian serine proteinase inhibitor
NGS: Next Generation Sequencing
NET: Neuroendocrine Tumors
PB: Pancreaticobiliary
PBC: Pancreaticobiliary cancers
RBC: Red Blood Cells
STR: Short Tandem Repeat
STROBE: Strengthening the Reporting of Observational Studies in Epidemiology
USG: Ultrasonography
USPSTF: United States Preventive Services Task Force
WBC: White Blood Cells;

## ETHICS APPROVAL AND CONSENT TO PARTICIPATE

All studies with human participants were approved by an Ethics Committee of the study sponsor and / or the participating site. The Institutional Ethics Committee (IEC) of Datar Cancer Genetics (DCG, sponsor) gave ethical approval for the TRUEBLOOD and RESOLUTE studies. The CTC-based Cancer Detection and Diagnosis Study was approved by the site ECs including Health Point Hospital (site), Krishna Institute of Medical Sciences (site) as well as an independent EC, viz., Royal Pune Independent Ethics Committee. All studies with human participants were performed in accordance with the Declaration of Helsinki and applicable regulatory guidelines. Written informed consent was obtained from all adult study participants prior to participation and sample collection.

## AVAILABILITY DATA AND MATERIALS

The datasets used and/or analysed during the current study are available from the corresponding author on reasonable request.

## AUTHOR CONTRIBUTIONS

The work reported in the paper has been performed by the authors, unless clearly specified in the text.

AG: Conceptualization, Methodology, Visualization, Writing – review and editing.

NR: Conceptualization, Methodology, Visualization, Writing – review and editing.

SL: Conceptualization, Methodology, Visualization, Writing – review and editing.

AdS: Conceptualization, Methodology, Visualization, Writing – review and editing.

RA: Conceptualization, Methodology, Visualization, Writing – review and editing.

DA: Conceptualization, Investigation, Methodology, Project Administration, Resources, Software, Supervision, Validation, Visualization, Writing – review and editing.

VD: Conceptualization, Funding acquisition, Project Administration, Resources, Supervision, Writing – review and editing.

AS: Data Curation, Formal Analysis, Investigation, Methodology, Software, Validation, Visualization, Writing – original draft.

DP: Conceptualization, Formal Analysis, Investigation, Methodology, Supervision, Visualization, Writing – original draft.

All authors have read the manuscript and agreed to its contents, the author list and its order as well as the above author contribution statements.

## Data Availability

All data produced in the present study are available upon reasonable request to the authors

## ACKNOWLEDGEMENTS

The authors express gratitude to the study participants as well as their caretakers / families. The authors acknowledge the contributions of the clinical staff at the participating sites and employees of the study sponsor for their contributions in managing various clinical, operational and laboratory aspects of the studies.

