## Supplementary Methods, Tables and Figures for "Liquid Biopsy for Detection of Pancreaticobiliary Cancers in Suspected Patients by Functional Enrichment and Immunofluorescent Profiling of Circulating Tumor Cells and their Clusters"

**Title**

**Table of Contents**

| **Section** | **Title** | **Page** |
| --- | --- | --- |
| **Supplementary Methods** | | |
| **SM1** | Markers | **2** |
| **SM2** | Antisera and Cell Lines | **2** |
| **SM3** | Enrichment of CTCs | **3** |
| **SM4** | Immunocytochemistry Profiling of CTCs | **3** |
| **SM5** | Marker Expression in Various Cell types | **4** |
| **SM6** | Marker Expression in PBC | **4** |
| **SM7** | PB-CTCs in Non-malignant Conditions | **4** |
| **SM8** | Analyte Stability | **4** |
| **SM9** | Linearity and Limit of Detection | **5** |
| **SM10** | Limit of Blank | **5** |
| **SM11** | Interference | **5** |
| **SM12** | Inter-Operator Agreement | **6** |
| **Supplementary Tables** | | |
| **ST1** | Marker Expression Study Cohort: PBC cases | **7** |
| **ST2** | Marker Expression Study Cohort: Non-malignant Conditions | **8** |
| **ST3** | Potentially Interfering Substances | **9** |
| **ST4** | Inter-Operator Concordance Study: Demographics | **10** |
| **ST5** | Inter-Operator Concordance Study: Findings | **11** |
| **ST6** | Case Control Clinical Study: Inclusion and Exclusion Criteria | **12** |
| **ST7** | Case Control Clinical Study: Participant Demographics | **13** |
| **ST8** | Case Control Clinical Study: Training Set Findings | **14** |
| **ST9** | Prospective Clinical Study: Participant Demographics | **15** |
| **ST10** | Prospective Clinical Study: Findings | **16** |
| **Supplementary Figures** | | |
| **SF1** | Marker Expression in Various Cell types. | **17** |
| **SF2** | PBC Stage and Marker Expression | **18** |
| **SF3** | Patient Age and Marker Expression | **19** |
| **SF4** | Patient Gender and Marker Expression | **20** |

**SUPPLEMENTARY METHODS**

**SM1. Markers**

The Test uses a multi-marker system which ensures the detection of Pancreaticobiliary Cancer (Adenocarcinoma) associated Circulating Tumor Cells (PB-CTCs) with high specificity, as malignant apoptosis resistant cells expressing (positive for) Carbohydrate Antigen 19-9 (CA19.9), Mammary Serine Protease Inhibitor (Maspin), Epithelial Cell Adhesion Molecule (EpCAM) and Cytokeratins (CK), and negative for the common leucocyte antigen (CD45). Carbohydrate Antigen 19-9 (CA19.9) plays a vital role in cell recognition processes and is used in the management of pancreatic, gallbladder and bile duct cancers. Mammary Serine Protease Inhibitor (Maspin) was originally reported to function as a tumor suppressor gene in epithelial cells, suppressing the ability of cancer cells to invade and metastasize to other tissues. Maspin protein is upregulated in pancreatic cancers but not in normal pancreatic tissue, thus aiding differentiation of pancreatic cancer from benign conditions. Maspin expression is also upregulated in gallbladder and bile duct cancers. Epithelial Cell Adhesion Molecule (EpCAM) are membrane antigens present on epithelial cells (and carcinomas) that function in cell adhesion. Cytokeratins (CK) are a family of cytoplasmic structural proteins expressed in epithelial tumors and CTCs. Common Leucocyte Antigen (CD45) as a negative marker serves to differentiate CTCs from CD45 positive haematolymphoid cells. CA19.9 and Maspin are evaluated during routine diagnostic histopathological evaluation (HPE) of Pancreaticobiliary tumor tissue. Co-expression of Maspin and CA19.9 is rare in organ systems outside of the Pancreas, Gallbladder, Bile Duct and thus ensures a high specificity of organ localisation.

**SM2. Antisera and Cell Lines**

The antisera used included recombinant human (RH) Anti CD326 IgG1-Vio 615 (Miltenyi Biotech), RH Anti-CK-IgG1-Vio 515 (Miltenyi Biotech), RH Anti-CD45-IgG1-APCVio 770 (Miltenyi Biotech), Mouse Anti-CA19.9 IgG1 (Dako), Mouse Anti-Maspin IgG1 (Dako) and Anti-Mouse Alexa Fluor 594 (Invitrogen). The reference cell lines PL45 and MOLT-3 were procured from American Type Culture Collection (ATCC). The purity of all cell lines was confirmed by periodic Short Tandem Repeat (STR) profiling. All cell lines were also periodically tested and verified to be Mycoplasma negativity.

**SM3. Enrichment of CTCs**

Aliquoted blood samples (5 mL) were processed for the enrichment of circulating tumor cells (CTCs) from white blood cells (WBC). Briefly, WBCs were isolated from whole blood via lysis of red blood cells (RBCs) followed by centrifugation. WBCs were resuspended in Phosphate Buffered Saline (PBS) and treated with a proprietary CTC enrichment medium (CEM) that induces cell death in all apoptosis-competent non-malignant (hemato-lymphoid, epithelial, and endothelial) cells, while malignant tumor derived cells (CTCs) survive due to apoptosis resistance. After treatment for 5 days at 37°C, surviving cells and cell clusters are harvested by gentle centrifugation (400 × *g*, 5 min, 4°C) and resuspended in PBS.

**SM4. Immunocytochemistry Profiling of CTCs**

Apoptosis reluctant cells enriched from 5 mL of blood were resuspended in 1500 μL 1x Phosphate Buffered Saline (PBS) and 100 μL aliquots of these cells were seeded into 15 wells. Cells were fixed with 4% Paraformaldehyde, Permeabilized with 0.3% Triton-X 100, and treated with 3% BSA (blocking). Cells were immunostained with each of the 3 separate Primary (1°) Ab cocktails for multiplexed analysis of the following combination of markers, (a) Anti-CK + Anti-CD45 + Anti-EpCAM, (b) Anti-CK + Anti-CD45 + Anti-CA19.9, (c) Anti-CK, Anti-CD45, Anti-Maspin. Samples for CA19.9 and Maspin were incubated with secondary (2°) anti-mouse Ab. PBS washes followed each Ab incubation step. Each marker combination was evaluated in 5 wells (333 μL × 5 = 1.67 mL equivalent of blood). Finally, cells were treated with 4’,6-Diamidino-2-phenylindole dihydrochloride (DAPI) for nuclear staining. Control samples (PL45 for EpCAM, CA19.9 and Maspin; MOLT3 for CD45) were included in each run. Samples were evaluated on the CellInsight High Content Screening (HCS) Platform to determine the Fluorescence Intensity (FI) for each marker. Marker expression was determined by the sequential excitation and acquisition of fluorescence signal. Marker positive cells are evaluated for other parameters including nucleated status, nucleus to cytoplasm (N:C) ratio and the localization of all markers to the expected cellular regions.

WBCs or viable cells isolated from benign or malignant tumors were resuspended in Phosphate Buffered Saline (PBS) solution and ICC profiled similarly as described above for CTCs.

**SM5.** **Marker Expression in Various Cell types**

Study: Reference cells (MOLT3, PL45), malignant tumor derived cells (M-TDC), PB-CTC, pooled CTCs from other cancer types (oCTC: Breast, Lung, Head and Neck, Cervix, Prostate and Ovary), WBC from patients with benign pancreaticobiliary conditions (B-WBC) or from healthy donors (HD-WBC) were immunostained to determine expression status of CK, EpCAM, CA19.9 and Maspin.

Findings: Higher expression of CK and EpCAM was seen in PL45, M-TDC, PB-CTCs and oCTCs, while expression of CA19.9 and Maspin was seen only in PL45, M-TDC and PB-CTC (**Supplementary Figure SF1**).

**SM6.** **Marker Expression in PBCs**

Study: FI for CA19.9, Maspin, EpCAM and CK were evaluated in subsets of PB-CTCs enriched from blood samples of patients with pancreatic, gallbladder and bile duct AD, stratified by age, gender or stage of cancer (**Supplementary Table ST1**).

Findings: There were no significant variations in expression any marker due to age-group, gender, primary organ or stage indicating that the test can detect PB-CTCs irrespective of these variables (**SF2 – SF4**).

**SM6. PB-CTCs in Non-malignant Conditions**

Study: To determine the Specificity of the Test to discern PBC from PBB, we evaluated blood samples from 20 individuals who were recently diagnosed with PBB. Samples were processed for CTC enrichment and ICC profiling as described above.

Findings: Among the blood samples from 20 known PBB cases (**ST2**), PB-CTCs were not detected in any of the samples.

**SM8. Analyte Stability**

Study: To determine the Analyte Stability, 4 × 5 mL of blood was collected from 5 known cases of PBC and processed at various time points including within 24 h (baseline), 24 h - 48 h, 48 h - 72 h and 72 h - 96 h after storage at 2°C - 8°C.

Findings: The recovery of marker positive cells at 0h – 24h (baseline) was initially normalized (considered as 100%). Then the recoveries at all other time points were represented as a fraction of the baseline. Based on this approach, ≥85% mean recovery of marker positive cells was observed up to 72 h in all patient samples. The minimal (<15%) reduction in recovery up to 72 h at 2-8°C indicated that samples received within 72 h could be considered pre-analytically equivalent and acceptable.

**SM9. Linearity and Limit of Detection**

Study: 168 × 5 mL aliquots of healthy donor blood were divided into 3 sets of 56 aliquots each (1 set per multiplexed marker combination). Aliquots in each set were spiked with (approximately) 1, 3, 5, 10, 20, 40 and 80 PL45 cells (8 replicates each) by serially dilutuing a PL45 master spike (10^5^ cells / 5 mL) prepared previously. The study also included 24 × 5 mL aliquots (3 sets × 8 replicates) of healthy donor blood samples which were not spiked. The recoveries of marker positive cells at each spike density were determined to ascertain proportionate (linear) response. The Limit of Detection (LoD) was determined from the replicate data of the 4 lowest level samples, i.e. 1, 3, 5 and 10 cells / 5 mL, as per the method described in CLSI EP17A2.

Findings: The recovery of marker positive cells showed linear characteristics in the evaluated range (1 – 80 cells / 5 mL) with no hook effect. The LoD was determined to be 2 cells / 5 mL for each of the 3 types of marker positive cells.

**SM10. Limit of Blank**

Study: The Limit of Blank (LoB) was determined from the 24 × 5 mL unspiked healthy donor blood samples in the Linearity study.

Findings: Since no CA19.9+, Maspin+ or EpCAM+ cells were detected in the unspiked samples (no false positives), the marker-wise and overall LoB was 0 cells / mL. The LoB study indicates the high specificity of the test, i.e., absence (low risk) of false positive findings in absence of analyte (marker positive cells).

**SM11. Interference**

Study: The performance characteristics of the Test were evaluated in presence of endogenous (serum markers) and exogenous (non-anticancer drugs) factors as possible interfering agents (**ST3**). Analytical grade molecules were used to prepare working stock solutions and immediately used for spiking studies. All drugs were evaluated at the reported Peak Plasma Concentrations (C_Max_), while serum markers were evaluated at concentrations that are considered as elevated. Peripheral blood (72 × 5 mL) from asymptomatic donors who had not taken any medication in the last 14 days was spiked with ~10 PL45 cells each. Each aliquot was used for detection of marker positive cells. Detection of either type of cells in all samples indicated absence of any interference from any of the above drugs.

Findings: The presence of non-anticancer drugs at medically relevant peak plasma concentrations (C_Max_) or the serum parameters evaluated did not significantly impact the recovery or detection of marker positive cells spiked into blood samples since recovery of marker positive cells was ≥ 80% in presence of any interfering agent (as compared to controls without any interfering agent). The test is expected to remain unaffected in presence of systemic treatment agents (drugs) and elevated serum parameters.

**SM12. Inter-Operator Agreement**

Study: Inter-operator (n = 2) agreements were determined using blood samples (10 mL) from a cohort of 75 individuals including 30 PBC cases, 15 PBB cases (**ST4**) and 30 asymptomatic individuals (no prior diagnosis or current suspicion of cancer). Samples were split into two aliquots of 5 mL each and one aliquot provided to each of two independent operators who were unaware of the clinical status of the samples. Samples were evaluated and reported as ‘C’ (cancer) or ‘B/H’ (benign/healthy). Test findings were compared with the clinical status to determine (a) concordance of marker findings with clinical status and (b) inter-operator concordance.

Findings: The sample-type wise findings are provided in **ST5.** The overall agreement (OA) was 96%, positive agreement (PA) was 90% (n = 30 cancers) and negative agreement (NA) was 100% (n = 15 benign conditions and 30 healthy individuals). Neither operator reported marker positive cells in samples from asymptomatic individuals or those with benign conditions. Operator 1 reported all cancer samples as positive while operator 2 reported 3 cancer samples as negative.

**SUPPLEMENTARY TABLES**

**ST1. Marker Expression Study Cohort: PBC cases.**

|  | **Pancreas** | **Gallbladder** | **Bile Duct** | **Overall** |
| --- | --- | --- | --- | --- |
| **Age Group**  *< 40 years*  *41 – 50 years*  *51 – 60 years*  *61 – 70 years*  *> 70 years* | *10*  *10*  *10*  *10*  *10* | *7*  *10*  *10*  *10*  *7* | *2*  *4*  *9*  *8*  *7* | ***19***  ***24***  ***29***  ***28***  ***24*** |
| **Gender**  *Female*  *Male* | *20*  *20* | *20*  *20* | *10*  *10* | ***50***  ***50*** |
| **Stage**  *Stage I*  *Stage II*  *Stage III*  *Stage IV* | *10*  *10*  *10*  *10* | *5*  *8*  *7*  *7* | *2*  *5*  *5*  *5* | ***17***  ***23***  ***22***  ***22*** |

**ST2. Marker Expression Study Cohort: Non-malignant Conditions.**

| **Parameter** | **Value** |
| --- | --- |
| **Age**  Median  Range | 48 years  17 – 81 years |
| **Gender**  Female  Male | 10  10 |
| **Diagnosis**  Benign Bile duct Stricture  Cholecystitis  Pancreatitis | 1  5  14 |

**ST3. Potentially Interfering Substances.** The endogenous factors represent the most commonly observed variables during blood pathology work-up. The exogenous factors selected for evaluation of interference represent the most prescribed non-anticancer medications in the US and Europe.

| **Endogenous Factors** | **Exogenous Factors** |
| --- | --- |
| Bilirubin,  Cholesterol,  Glucose,  Haemoglobin,  Uric Acid | Levothyroxine,  Lisinopril,  Atorvastatin,  Metformin,  Amlodipine,  Metoprolol,  Omeprazole,  Albuterol,  Ranitidine,  Azithromycin,  Paracetamol,  Aspirin,  Loperamide,  Dextromethorphan,  Cortisone,  Ulipristal Acetate,  Sildenafil Citrate,  EDTA |

**ST4.** **Inter-Operator Concordance Study: Participant Demographics.**

|  | **Pancreas** | **Gallbladder** | **Bile Duct** | **Combined** |
| --- | --- | --- | --- | --- |
| **Cancers** | | | | |
| **N =** | 12 | 13 | 5 | 30 |
| **Age (years)**  Median  Range | 62  (23 – 79) | 53  (19 – 66) | 46  (36 – 72) | 57  (19 – 79) |
| **Gender**  Male  Female | 5  7 | 4  9 | 3  2 | 12  18 |
| **Stage**  Stage I  Stage II  Stage III  Stage IV | 5  1  1  5 | 1  1  2  9 | 1  1  2  1 | 7  3  5  15 |
| **Benign conditions** | | | | |
| **N =** | 13 | 2 | - | 15 |
| **Age (years)**  Median  Range | 36  (6 – 71) | 62  (62 – 62) | -  - | 38  (6 – 71) |
| **Gender**  Male  Female | 7  6 | 2  - | -  - | 9  6 |
| **Healthy (asymptomatic) adults** | | | | |
| **N =** | - | - | - | 30 |
| **Age (years)**  Median  Range | -  - | -  - | -  - | 38  (24 – 56) |
| **Gender**  Male  Female | -  - | -  - | -  - | 30  - |

**ST5. Inter-Operator Concordance Study: Findings**

| **Clinical status** | | | **Classification** | | | |
| --- | --- | --- | --- | --- | --- | --- |
|  |  |  | **Operator 1** | | **Operator 2** | |
| **Sample Type** | **Type** | **N =** | **C** | **B/H** | **C** | **B/H** |
| **All Healthy** | **H** | **30** | **-** | **30** | **-** | **30** |
| **All Benign** | **B** | **15** | **-** | **15** | **-** | **15** |
| *Cholecystitis* | *B* | *2* | *-* | *2* | *-* | *2* |
| *Pancreatitis* | *B* | *11* | *-* | *11* | *-* | *11* |
| *Pancreas IMT* | *B* | *2* | *-* | *2* | *-* | *2* |
| **All Cancers** | **C** | **30** | **30** | **-** | **27** | **3** |
| *Pancreas AD*  *Stage 1*  *Stage 2*  *Stage 3*  *Stage 4* | *C* | *12*  *5*  *1*  *1*  *5* | *12*  *5*  *1*  *1*  *5* | *-*  *-*  *-*  *-*  *-* | *9*  *4*  *-*  *-*  *5* | *3*  *1*  *1*  *1*  *-* |
| *Gallbladder AD*  *Stage 1*  *Stage 2*  *Stage 3*  *Stage 4* | *C* | *13*  *1*  *1*  *2*  *9* | *13*  *1*  *1*  *2*  *9* | *-*  *-*  *-*  *-*  *-* | *13*  *1*  *1*  *2*  *9* | *-*  *-*  *-*  *-*  *-* |
| *Bile Duct AD*  *Stage 1*  *Stage 2*  *Stage 3*  *Stage 4* | *C* | *5*  *1*  *1*  *2*  *1* | *5*  *1*  *1*  *2*  *1* | *-*  *-*  *-*  *-*  *-* | *5*  *1*  *1*  *2*  *1* | *-*  *-*  *-*  *-*  *-* |
| *B: benign; H: healthy; C: cancer; IMT: Inflammatory myofibroblastic tumor (benign)* | | | | | | |

**ST6. Case Control Clinical Study: Inclusion and Exclusion Criteria.**

|  | **Cancer Cases** | **Healthy Adults** |
| --- | --- | --- |
| **Inclusion**  (all) | Adult males and females,  Recent diagnosis of PBC,  Treatment Naïve | Adult males and females,  Aged 40 years and above,  No history of cancer diagnosis,  No present suspicion of cancer,  Normal serum CA19.9,  Normal USG (A+P),  Asymptomatic. |
| **Exclusion**  (any) | Other cancers,  Received anticancer treatments | Prior diagnosis of cancer,  Currently suspected of cancer,  Elevated serum CA19.9,  Suspicious findings on USG |

**ST7. Case Control Clinical Study: Participant Demographics**. This study included 5 mL blood samples from 188 recently diagnosed and therapy naïve PBC cases (cases) and 172 asymptomatic individuals (controls).

| **Parameter** | **Cancer Cases** | | | | **Healthy Adults** |
| --- | --- | --- | --- | --- | --- |
|  | **Pancreas** | **Gallbladder** | **Bile Duct** | **Overall** |  |
| **N =** | **117** | **44** | **27** | **188** | **172** |
| **Age (years)**  Median  Range | 60  13 - 83 | 55  19 - 74 | 60  36 - 81 | 60  13 - 83 | 49  40 - 82 |
| **Gender**  Female  Male | 48  69 | 27  17 | 8  19 | 83  105 | 70  102 |
| **Stage**  Stage I  Stage II  Stage III  Stage IV | 36  41  20  20 | 11  11  10  12 | 3  9  7  8 | 50  61  37  40 |  |

**ST8. Case Control Clinical Study: Training Set Findings.** The Training Set of 252 samples included 132 PBC cases and 120 asymptomatic individuals. All 120 samples from asymptomatic individuals were negative for PB-CTCs. Among the 132 samples from cancer patients, 126 were positive for PB-CTCs and 6 were negative.

| **Sample Type** | **Negative** | **Positive** |
| --- | --- | --- |
| **Asymptomatic (n = 120)** | **120 (100.0%)** | **0 (0.0%)** |
| **All Cancers (n = 132)**  *Stage I (n = 35)*  *Stage II (n = 43)*  *Stage III (n = 26)*  *Stage IV (n = 28)* | **6 (4.5%)**  *6 (17.1%)*  *-*  *-*  *-* | **126 (95.5%)**  *29 (82.9%)*  *43 (100.0%)*  *26 (100.0%)*  *28 (100.0%)* |
| ***Cancer-wise*** | | |
| **Pancreas (n = 82)**  *Stage I (n = 25)*  *Stage II (n = 29)*  *Stage III (n = 14)*  *Stage IV (n = 14)*  **Gallbladder (n = 31)**  *Stage I (n = 8)*  *Stage II (n = 8)*  *Stage III (n = 7)*  *Stage IV (n = 8)*  **Bile Duct (n = 19)**  *Stage I (n = 2)*  *Stage II (n = 6)*  *Stage III (n = 5)*  *Stage IV (n = 6)* | **4 (4.9%)**  *4 (16.0%)*  *-*  *-*  *-*  **1 (3.2%)**  *1 (12.5%)*  *-*  *-*  *-*  **1 (5.3%)**  *1 (50.0%)*  *-*  *-*  *-* | **78 (95.1%)**  *21 (84.0%)*  *29 (100.0%)*  *14 (100.0%)*  *14 (100.0%)*  **30 (96.8%)**  *7 (87.5%)*  *8 (100.0%)*  *7 (100.0%)*  *8 (100.0%)*  **18 (94.7%)**  *1 (50.0%)*  *6 (100.0%)*  *5 (100.0%)*  *6 (100.0%)* |

**ST9. Prospective Clinical Study: Participant Demographics.** The prospective clinical study included 88 individuals suspected of PBC, who were advised a biopsy for diagnosis.

| **Parameter** | **Post HPE Status of all samples** | | | | |
| --- | --- | --- | --- | --- | --- |
|  | ***Cancers*** | | | | **Benign** |
|  | ***Pancreas*** | ***Gallbladder*** | ***Bile Duct*** | ***Combined*** |  |
| **N =** | 23 | 19 | 7 | 49 | 39 |
| **Age (years)**  Median  Range | 58  33 – 70 | 45  31 – 62 | 60  29 - 78 | 56  29 - 78 | 38  21 - 81 |
| **Gender**  Female  Male | 5  18 | 13  6 | 6  1 | 24  25 | 21  18 |
| **Cancer Stage^#^**  Stage I  Stage II  Stage III  Stage IV | 12  2  4  5 | 9  2  2  6 | -  1  4  2 | 21  5  10  13 |  |
| **Benign Condition^#^**  Bile Duct Stricture  Cholecystitis  Cholelithiasis  Pancreatic BIMT*  Pancreatitis  Heterotypic Pancreas  Cystic Fibrosis of Pancreas |  |  |  |  | 2  4  5  2  24  1  1 |
| **Benign Inflammatory myofibroblastic tumor.* | | | | | |

**ST10. Prospective Clinical Study: Findings.** Based on observed marker expression profile in the 88 samples, 50 were classified Positive and 38 were classified Negative.

| **Sample Type** | **Negative** | **Positive** |
| --- | --- | --- |
| **Benign (n = 39)** | **36 (92.3%)** | **3 (7.7%)** |
| **All Cancers (n = 49)**  *Stage I (n = 21)*  *Stage II (n = 5)*  *Stage III (n = 10)*  *Stage IV (n = 13)* | **2 (4.1%)**  *2 (9.5%)*  *-*  *-*  *-* | **47 (95.9%)**  *19 (90.5%)*  *5 (100.0%)*  *10 (100.0%)*  *13 (100.0%)* |
| ***Cancer-wise*** | | |
| **Pancreas (n = 23)**  *Stage I (n = 12)*  *Stage II (n = 2)*  *Stage III (n = 4)*  *Stage IV (n = 5)*  **Gallbladder (n = 19)**  *Stage I (n = 9)*  *Stage II (n = 2)*  *Stage III (n = 2)*  *Stage IV (n = 6)*  **Bile Duct (n = 7)**  *Stage I (n = 0)*  *Stage II (n = 1)*  *Stage III (n = 4)*  *Stage IV (n = 2)* | **1 (4.3%)**  *1 (8.3%)*  *-*  *-*  *-*  **1 (5.3%)**  *1 (11.1%)*  *-*  *-*  *-*  **-**  *-*  *-*  *-*  *-* | **22 (95.7%)**  *11 (91.7%)*  *2 (100.0%)*  *4 (100.0%)*  *5 (100.0%)*  **18 (94.7%)**  *8 (88.9%)*  *2 (100.0%)*  *2 (100.0%)*  *6 (100.0%)*  **7 (100.0%)**  *-*  *1 (100.0%)*  *4 (100.0%)*  *2 (100.0%)* |

**SUPPLEMENTARY FIGURES**

**SF1. Marker Expression in Various Cell types.** The expression (FI: fluorescence intensity) of CK, EpCAM, CA19.9 and Maspin were evaluated on various cell types.

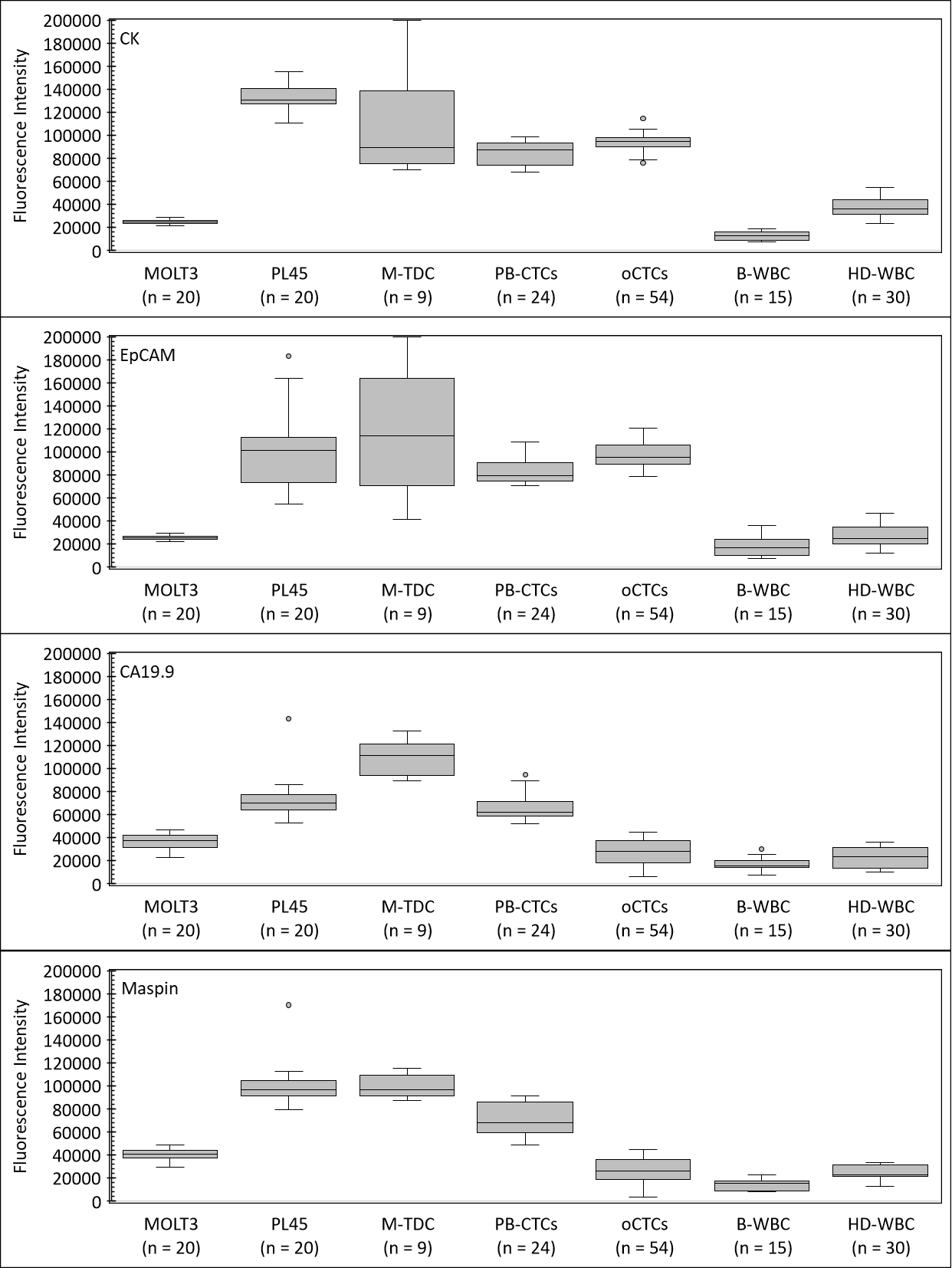

**SF2. PBC Stage and Marker Expression.** (P) Pancreas, (GB) Gallbladder, (BD) Bile duct, (St) Stage.

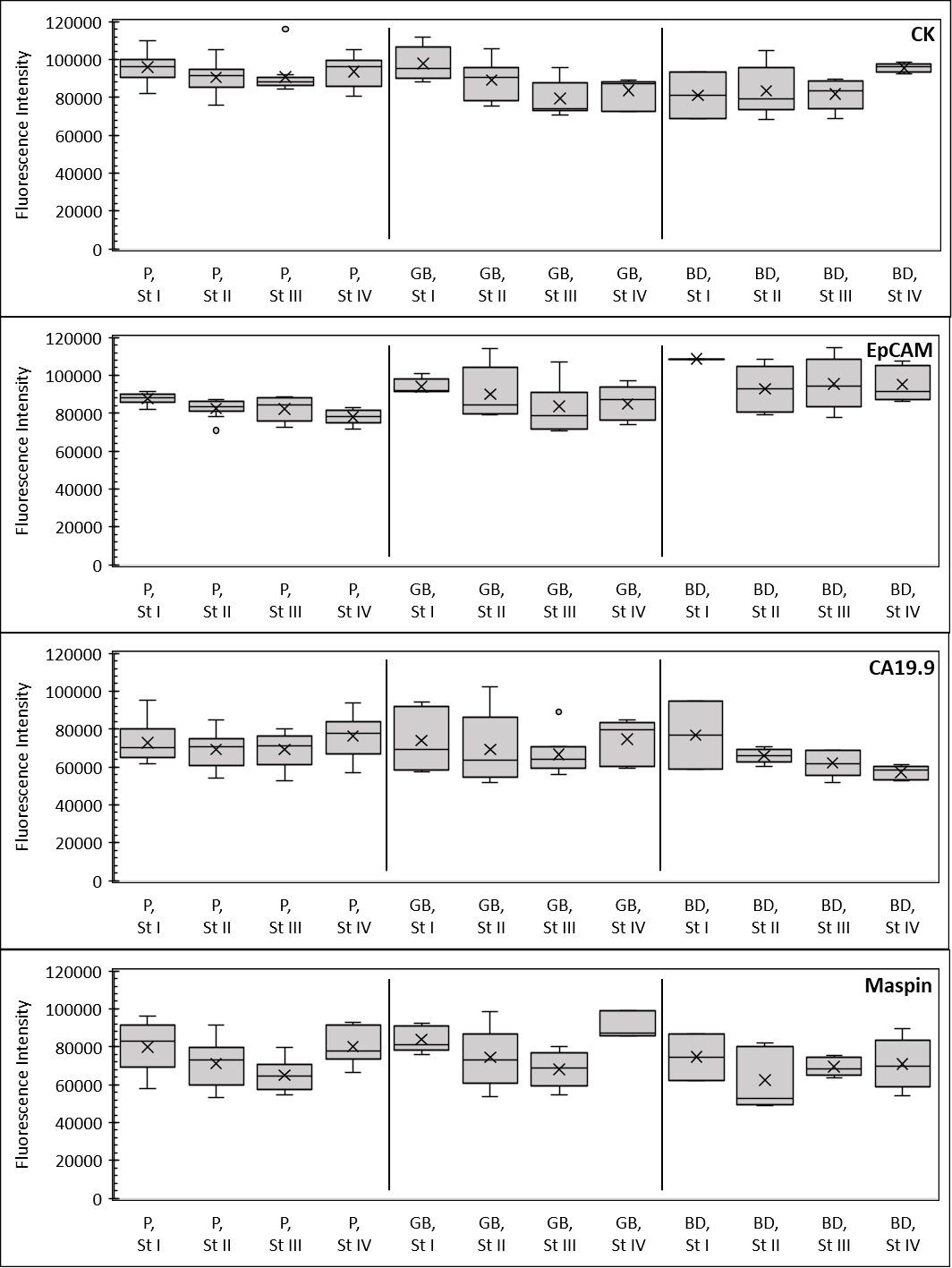

**SF3. Patient Age and Marker Expression.** (P) Pancreas, (GB) Gallbladder, (BD) Bile duct. Age ranges are indicated in years.

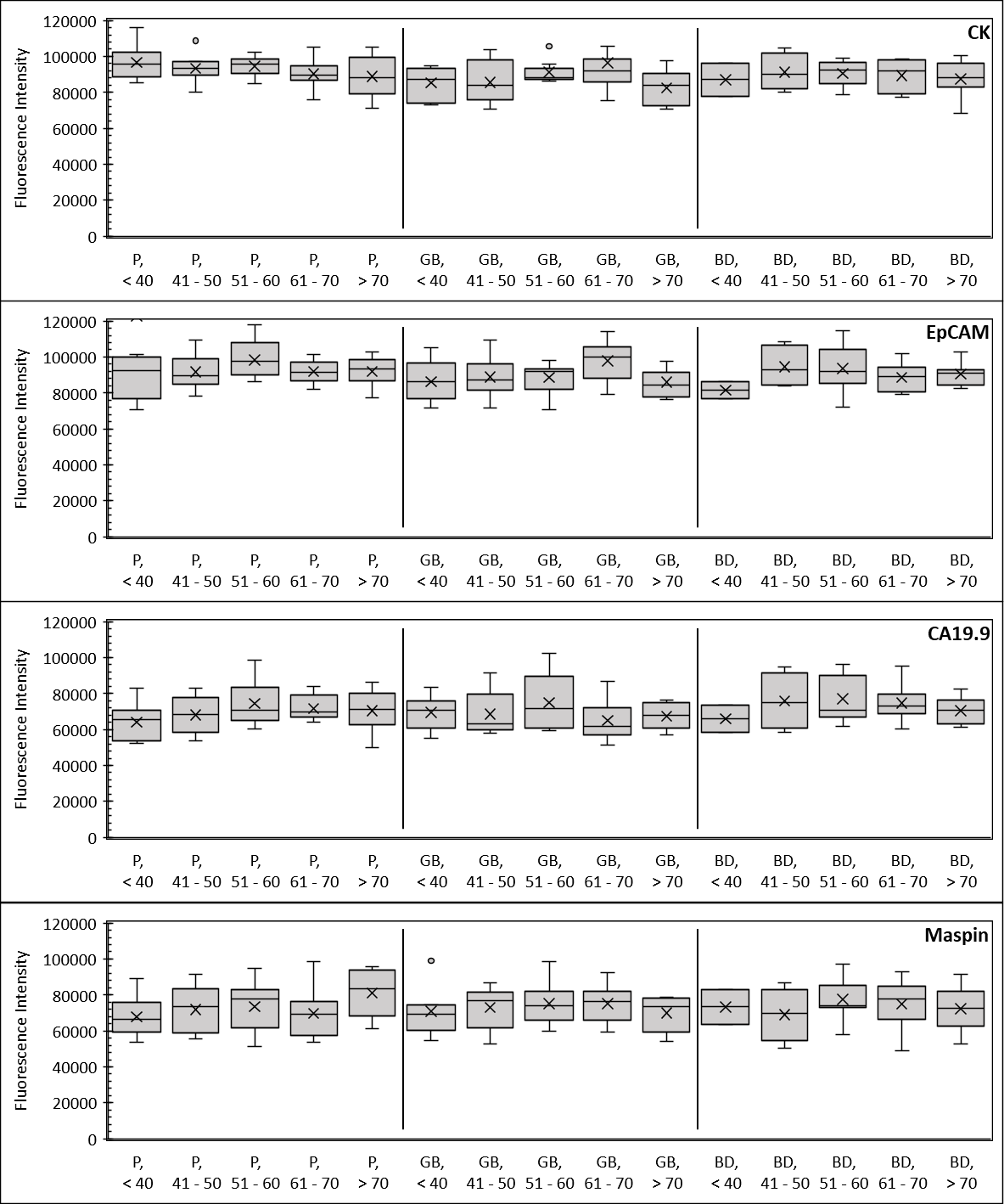

**SF4. Patient Gender and Marker Expression.** (P) Pancreas, (GB) Gallbladder, (BD) Bile duct, (F) Female, (M) Male.

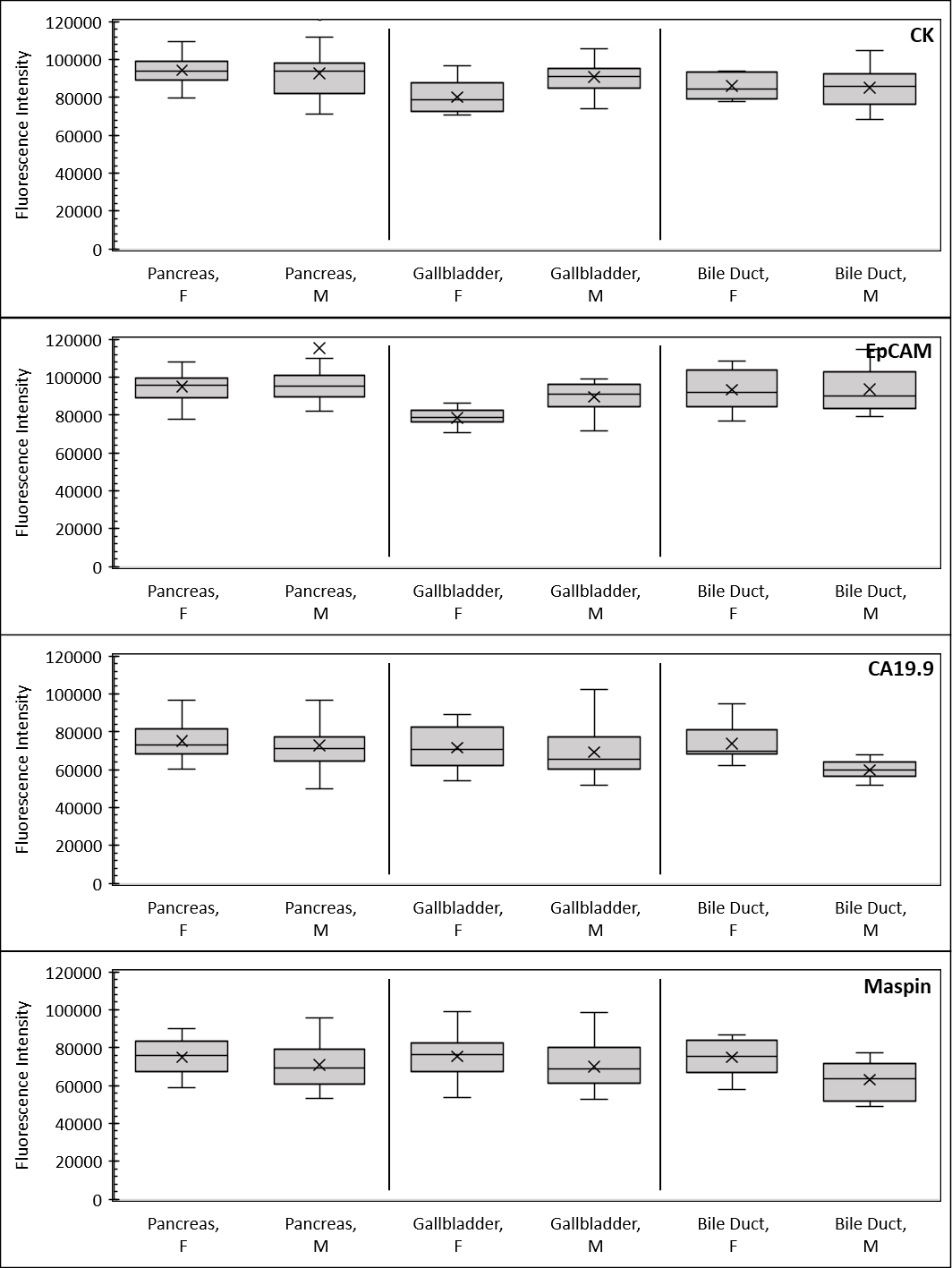
